# Randomised study of a new inline respiratory function monitor (Juno) to improve mask seal and delivered ventilation with neonatal manikins

**DOI:** 10.1101/2023.09.12.23295396

**Authors:** Mark Tracy, Murray Hinder, Stephanie Morakeas, Krista Lowe, Archana Priyadarshi, Matthew Crott, Matthew Boustred, Mihaela Culcer

## Abstract

**Background:** Respiratory function monitors (RFM) have been used extensively in manikin and infant studies yet have not become the standard of training. We report the outcomes of a new portable, lightweight resuscitation monitor (Juno) designed to show mask leak and deflation tidal volume to assist in positive pressure ventilation (PPV) competency training using manikins.

**Methods:** Two leak-free manikins (Preterm and Term) were used. Participants provided positive pressure ventilation to manikins using two randomized devices self-inflating bag (SIB) and T-piece resuscitator (TPR) with Juno display initially blinded then unblinded in four 90-second paired sequences, aiming for adequate chest wall rise and target minimal mask leak with appropriate target delivered volume when using the monitor.

**Results:** 49 experienced neonatal staff delivered 15,569 inflations to the term manikin and 14,580 inflations to the preterm. Comparing blinded to unblinded RFM display there were significant reductions in all groups in the number of inflations out of target range volumes (Preterm: SIB 22.6% to 6.6%, TPR: 7.1% to 4.2% and Term: SIB 54.8% to 37.8%, TPR 67.2 to 63.8%). The percentage of mask leak inflations >60% was reduced in Preterm: SIB 20.7% to 7.2%, TPR 23.4% to 7.4% and in Term: SIB 8.7% to 3.6%, TPR 23.5% to 6.2%).

**Conclusions:** Using the Juno monitor during simulated resuscitation significantly improved mask leak and delivered ventilation amongst otherwise experienced staff, using both preterm and term manikins. The Juno is a novel RFM that may assist in teaching and self-assessment of resuscitation technique.

**Key messages:** *What is already known on this topic:*

- Mask leak is common, can be large in magnitude and produce ineffective ventilation.
- Excessive tidal volumes may injure vulnerable preterm lungs and brain.
- The optimum use of RFMs used in manikin training is not determined.

*What this study adds:*

- Mask leaks in excess of 60 % significantly reduces delivered tidal volumes.
- Optimising mask seal reduces high leak inflations in manikin models.
- Optimising mask seal alone with TPR improves targeted volume delivery in manikin models.

*How this study might affect research, practice or policy:*

- Inline RFM monitors with simple GUI can improve the delivery of resuscitation training at all levels of skill.
- Further research to guide policy how often to train to retain skills is needed.
- Further research will determine the effectiveness of this RFM as a tool for self-directed learning in rural or remote settings.

## INTRODUCTION

Effective ventilation is one of the critical techniques of newborn resuscitation. An essential characteristic is achieving adequate mask seal to deliver appropriate lung inflation during PPV. ^1 2^ This skill is required by a wide range of health practitioners and depends on repetitive training. As well as experienced neonatologists, first responders may include midwives, nurses and doctors not based in NICU. The retention of resuscitation skills relies on practice and assessment using manikins, typically with no biofeedback on delivered ventilation, mask leak or functionality of resuscitation device. In many circumstances manikins used to assess training skills may be limited by the range of size, structural characteristics (how much force is required to achieve an adequate seal) and functional system compliance. Damaged manikins can produce unrecognized and unintended internal leaks. These factors lead practitioners to apply more inflation pressure or mask force to achieve a given chest wall movement. Our research has shown that many brands of resuscitation devices in leak-free settings can fail to deliver adequate ventilation despite compliance with current international standards. ^3^

Respiratory function monitors have been used in many manikin and human infant studies to quantitate mask seal as a prerequisite to appropriate delivered tidal volumes. ^4 5^ Most studies have used either the commercial Philips NM3 or the Acutronics Florian RFM or research RFMs that inform current guidelines on RFM use during newborn resuscitation. ^6-10^ They are expensive, heavy and complex monitors that require expert knowledge to use and are no longer commercially available. There is a need for a new class of monitor for resuscitation that enables the practitioner to access real-time data in an optimal display of ventilation performance and immediately correct technique if there is an excessive leak or inflation volumes.

This paper describes the assessment of a novel resuscitation monitor, the “Juno” monitor built by ResusRight^11^ and used in a manikin study. The monitor is a miniaturised battery driven RFM small enough to fit between the patient interface (face mask, laryngeal mask or endotracheal tube) and resuscitation device. The LED screen has been designed to be in the line of sight with the manikin or baby without obscuring rise and fall of the chest. The Graphical User Interface (GUI) is an LED screen that displays easily interpreted icons (three baby sizes) and a ‘traffic light’ system for mask leak range. Thus, critical information relating to deflation/expired tidal volume (tve) in mL according to the size of the infant being resuscitated and mask leak can be interpreted (Figure 1& Figure 3s).

**Figure 1:**
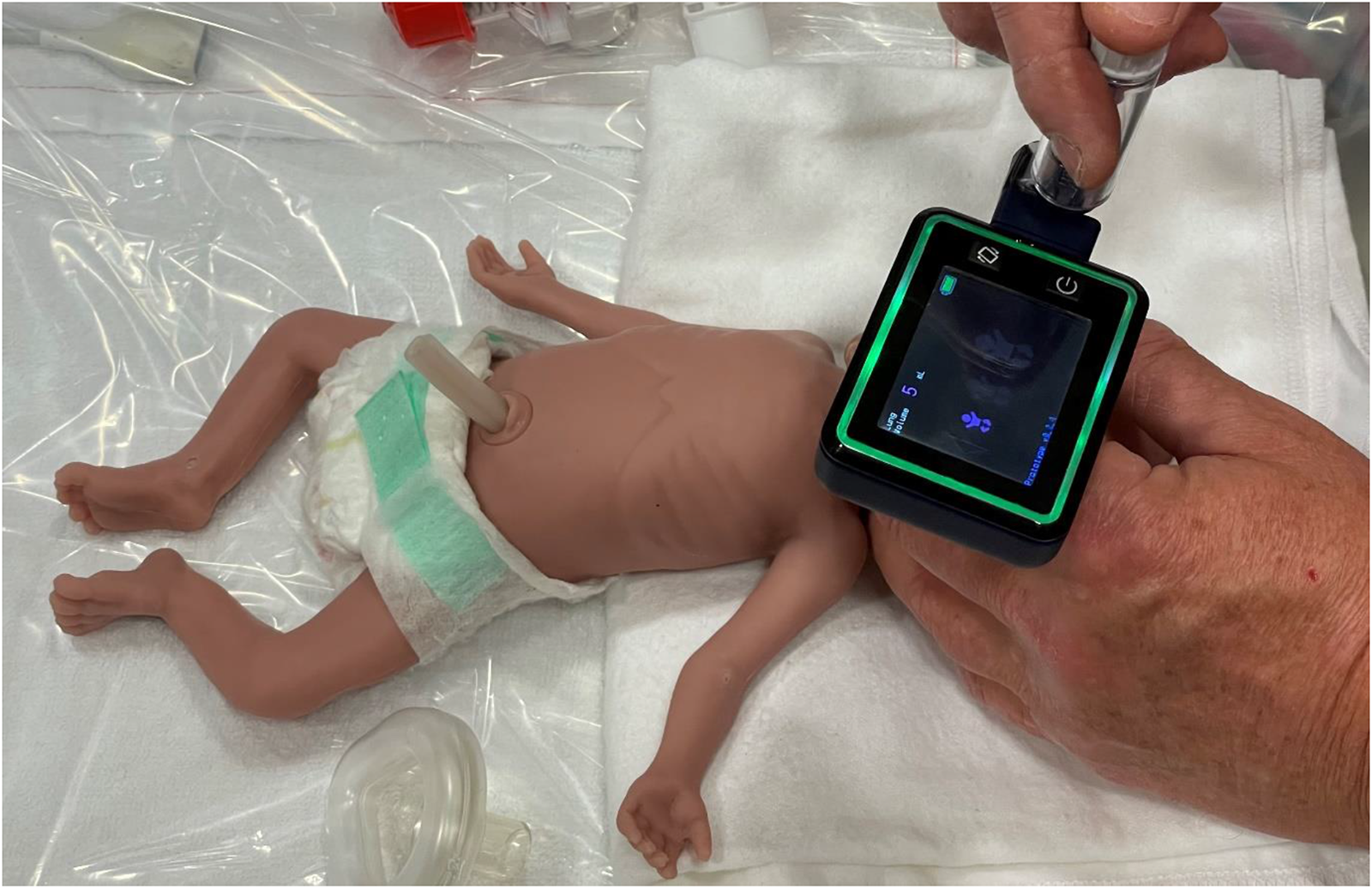
The Juno monitor studied pre-commercial prototype

We aimed to compare the mask leak and delivered lung volumes using the Juno resuscitation monitor with the display screen blinded and unblinded, using two different-sized manikins (Laerdal Premature Anne and the Laerdal ALS infant manikins) and two different inflation devices: Ambu SIB; and Fisher & Paykel Neopuff TPR. Our null hypothesis was that there will be no difference in the targeted delivered volumes, and mask leak applied to the manikins with or without the Juno monitor display unblinded or blinded. Ethics approval was obtained from Sydney Children’s Hospital Network ethics committee(#2019/ETH13537).

## MATERIALS AND METHODS

### Setting

Staff of a newborn intensive care nursery at a major metropolitan teaching hospital (Westmead Sydney, Australia) were invited to participate. 49 experienced NICU staff (nurses, nurse practitioners, doctors: junior & senior) consented to participate. All had previously received extensive training in neonatal resuscitation, demonstrating proficiency annually in the locally run neonatal intensive care unit resuscitation course. This course uses the American Academy of Pediatrics neonatal resuscitation program^12^. The mask hold taught for a single person is the two-point top. ^1^ Importantly, all participants had received repeated training and exposure to the Juno resuscitation monitor, were familiar with volume targeting and mask leak indication during PPV and were assessed to be confident in using the monitor before the study.

### Manikins

We used the Laerdal ALS trainer infant (#08003040) and the Premature Anne (#290-00050) manikins. Both were tested for leaks and found to be leak free. The ALS infant manikin has a hinged mandible allowing a realistic jaw thrust and a closed system with lung and stomach bags, the esophageal tube was blocked for this study. Static compliance was measured at 2.4 mL/cm H_2_O. The Premature Anne approximates a 25-week pre-term manikin with measured static compliance of 0.6mL/cmH_2_O.

### Ventilation devices

A new disposable Ambu Spur II Infant Self Inflating Bag (volume 220mL) with reservoir bag (#335 102 000), Ambu disposable Peep valve 20 (#199 102 001) and Ambu manometer (#322 003 000) attached (Ambu A/S Ballerup, Denmark) was used with each participant. No auxiliary gas inflow was used. The T-piece resuscitator (TPR) used was a Neopuff infant (#RD900 Fisher & Paykel Health Care New Zealand) with gas inflow set to 10 LPM. The mask used were Ambu triangular disposable face mask (#000 252 951) for term and Fisher & Paykel Neonatal resuscitation 35mm mask preterm (#RD803).

### The resuscitation monitor (Juno)

The Juno is a lightweight (85g), inline, battery-driven (run-time 5 hours) RFM. The Juno monitor studied was a pre-commercial prototype (SW V0.2.4) displaying: mask leak grouped by a traffic light LED panel (green reflecting leak from 0 to 29.9%, orange from 30 to 59.9% and red ≥ 60 to 100%); deflation tidal volume in mL as well as baby range icons (small 2.5 - 9.9mL, medium 10 – 24.9mL and large 25 – 50mL) and inflation rate per minute (IPM). Displayed data is updated for each inflation in real time. Small and large baby volume icons change red to indicate low (<2.5mL) and excessive (≥ 50mL) tve, and a no-breath icon indicates after 5 seconds of no airflow detected (Figure 4s). The monitor is situated in the line of sight of the chest wall thus enabling the resuscitator to visualize the monitor and the chest wall at the same time.

The Juno monitor uses a thermal mass flow pneumotach to detect flow in the cuvette and has inbuilt memory storing inflation-by-inflation data. The Juno monitor does not measure pressure, only flow that integrates into volumes. It was extensively validated with traceable reference testing systems. Volume reference using calibrated precision syringes (Hans Rudolph series 5520) ^13^ +/- 0.05mL and, calibrated flow reference testing +/- 1.75% of reading or +/- 0.05 sL/min (IMT Analytics AG Flow Analyser PF-300) ^14^. Juno was found to be well within stated accuracy +/-8% of volume readings. Data stored is time, inflation tidal volume(tvi), tve, inflation time(ti) and deflation time(te). The Juno is currently approved for use in neonatal resuscitation training in Australia and Europe and is not approved for clinical use on humans.

### Data Collection

Two separate data collection sessions were carried out, one for each manikin size. Participants were randomized for starting resuscitation device (TPR or SIB), Juno device was in situ for all combinations and the Juno display was initially blinded out for each resuscitation device.

### Participant Instructions

With the Juno display blinded the task was to provide 90 seconds of mask PPV to the manikin to achieve adequate chest wall rise and use a rate between 40 to 60 inflations per NRP guidelines^12^. For the preterm manikin, the target pressures were 20 cmH_2_O Peak Inspiratory Pressure (PIP) and PEEP 5 cmH_2_O (SIB, PEEP preset, and PIP targeted by manometer; TPR preset to 20/5). For the term manikin, target pressures were 25 cmH_2_O PIP and PEEP 5 cmH_2_O (SIB: PEEP preset, and PIP targeted by manometer; TPR: preset to 25/5).

With the Juno display unblinded the task was to provide 90 seconds of PPV to the manikin to achieve adequate chest wall rise and use a rate between 40 to 60 inflations per minute. Participants were asked to use the mask leak visual indicators (Figure 1) to optimize the mask seal by adjusting their mask hold technique if necessary (if the leak indicator was red or orange) and when using SIB target appropriate PIP to achieve targeted volumes, using either the actual tve volume display (preterm 4-6mLs, Term 25-30mLs) or the baby icon volume range (small baby 2.5 to 10 mLs or large baby icon baby 25 to 50 mLs). With TPR use, participants were asked to minimize mask leak only (they did not adjust TPR PIP to target tve, PIP remained at the preset value). Participants had a two-minute rest between changes in display status or resuscitation device used. Data were downloaded via USB-C cable. Less experienced staff were encouraged to use the icons to determine volumes.

### Data analysis

Analysis was conducted using Stata (V.17 MP). The measured test lung parameters were tvi, tve, inflation, and deflation time. Mask leak percentage was calculated using the formula ((tvi-tve)/tvi) X100. A-priori the first two inflations (mask applied) and the last inflation (mask released) were removed. Inflations during mask seal adjustments typically resulted in tvi/tve/ti/te or te outside meaningful ranges and were removed. These reflected mask seal adjustment periods (prior validation bench assessments). Mask leaks with negative values between -15 and 0 % were examined and re-coded to 0 leak %. Negative leaks < -15% were discarded. A total of 1,452 (8.49%) of inflations for term manikin were removed and 2,238 (13.8%) for preterm. Raw data were examined for distributional characteristics. We used univariate logistic regression to estimate odds ratios (ORs) with 95% CIs and Pearson χ^2^ tests to calculate p values to compare proportions between the study groups for the main dichotomous outcomes. For non-normally distributed data we used non-parametric bootstrap median regression with 95 CIs to infer the observed significance of the effects (Median deflation tidal volumes by leak group). Analysis of variance (ANOVA) for repeated measures was used to determine differences in predicted means between devices and screen unblinded or not adjusting for individuals. ANOVA was reported with p values adjusted F test using Box’s conservative epsilon, p values of <0.05 were considered statistically significant.

## RESULTS

There was a total of 14,580 inflations recorded with the preterm manikin (SIB 7,356 (50.4%) NP 7,224 (49.6%)) and 15,659 inflations with the term manikin (SIB 8,160 (52%) NP 7,499 (48%)). As mask leak increased by groups according to leak, the delivered inflation volume decreased significantly in both manikin models. (Figure 2 and Table 1) P<0.001.

**Table 1:** Inflation count and percent of deflations in target volume range.

|  | TPR |  | SIB |  | Total |
| --- | --- | --- | --- | --- | --- |
|  | blinded | unblinded | blinded | unblinded |  |
| **Term** |  |  |  |  |  |
| In range 25-40mL | 1,226(32.8%)** | 1,359(36.2%)** | 1,854(45.2%)* | 2,547(62.7%)* |  |
| Out of range | 2,514(67.2%) | 2,400(63.8%) | 2,245(54.8%) | 1,514(37.8%) |  |
| Inflations | 3,740 | 3,759 | 4,099 | 4,061 | 15,659 |
| **Preterm** |  |  |  |  |  |
| In range 2.5-10mL | 3,271(93%)** | 3,546(95.8%)** | 2,548(77.4%)* | 3,796(93.4%)* |  |
| Out of range | 250(7.1%) | 157(4.2%) | 743(22.6%) | 269(6.6%) |  |
| Inflations | 3,521 | 3,703 | 3,291 | 4,065 | 14,580 |
| Total Inflations |  |  |  |  | 30,239 |

\* $p<0.001$ , \*\* $p=0.002$ Chi<sup>2</sup>

**Figure 2:**
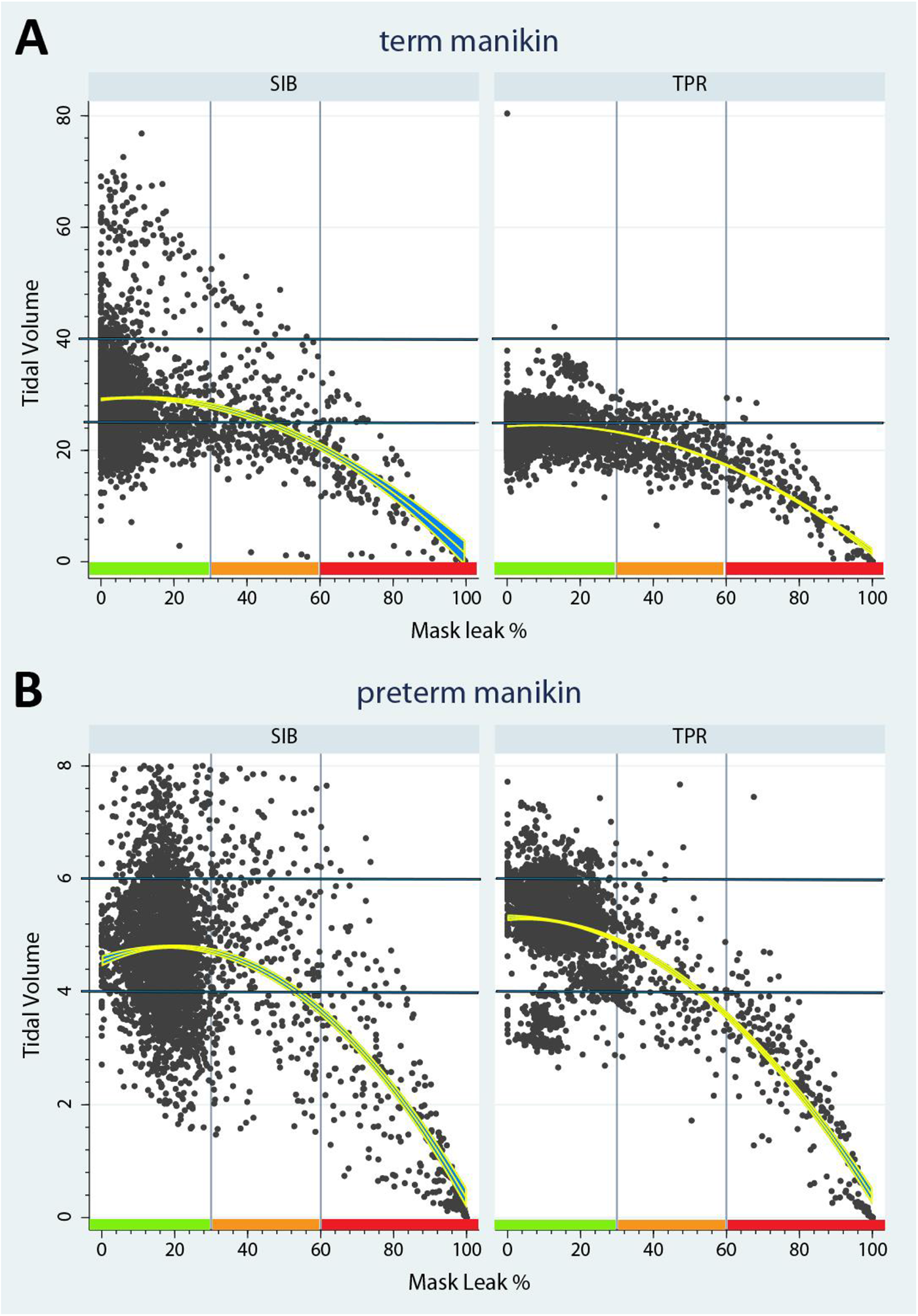
Term Manikin (A) and Preterm (B) scatter plots of inflations with Juno display unblinded. By target volume ranges 4-6 mLs and 25-50 mLs, and percent Leak category: Green 0 – 29.9%, Orange 30-59.9%, Red 60 -100% Leak. Regression line is quadratic with 95% CI.

More inflations were delivered in the target range when the Juno monitor was unblinded for both devices and manikins p<0.005 with a more pronounced difference with the SIB. With the monitor unblinded there was a significant reduction in the number of inflations with mask leak >60% (term manikin: TPR 23.5% blinded vs 6.2% unblinded (OR -4.9) and with SIB 8.7% blinded vs 3.6% unblinded (OR -3.1)), (preterm manikin: TPR 23.5% blinded vs 7.4% unblinded (OR -1.5) and SIB 20.8% blinded vs 7.2% unblinded (OR -1.5)) p<0.001 (Table 2). There was a significant and clinically meaningful reduction in predicted mean leak values in term manikin and TPR from predicted means of 29.9% blacked to 16.4% and with SIB 15.8% blinded ed to 10.5% unblinded (Table 3). Similar significant reductions were seen with predicted mask leaks of TPR blinded mean 33% reduced to 18.3% and with SIB 38% reduced to 22.6% p<0.001.

**Table 2:** Inflation count and percentages by leak display category, device and Juno monitor displayed. Adjusted Odd Ratio referent group green mask leak with 95% CI.

|  | TPR |  |  | SIB |  |  |
| --- | --- | --- | --- | --- | --- | --- |
|  | Blinded | Unblinded | Adjusted OR | Blinded | Unblinded | Adjusted OR |
| **Term** |  |  |  |  |  |  |
| Green | 2,376 (63.5%) | 3151 (83.8%) | Ref 95% Ci | 3,403 (83.0%) | 3,594 (88.5%) | Ref 95% Ci |
| Orange | 484 (12.9%) | 374 (9.9%) | -1.9(-2.2 to -1.7) | 338 (8.3%) | 319 (7.8%) | -0.9 (-1.1 to -0.7) |
| Red | 880 (23.5%) | 234 (6.2%)* | -4.9 (-5.6 to -4.12) | 358 (8.7%) | 148 (3.6%)* | -3.1 (-3.5 to -2.8) |
| Inflations | 3,740 | 3,759 |  | 4,099 | 4,061 |  |
| **Preterm** |  |  |  |  |  |  |
| Green | 2,284 (64.9%) | 3,215 (86.8%) | Ref 95% Ci | 1,861(56.5%) | 3,435(84.5%) | Ref 95% Ci |
| Orange | 411 (11.7%) | 215 (5.8%) | -1.0(-1.1 to -0.8) | 747 (22.7%) | 338 (8.3%) | -1.4(-1.6 - -1.3) |
| Red | 826 (23.5%) | 273 (7.4%) | -1.5(-1.6 to -1.3) | 683 (20.8%) | 292 (7.2%) | -1.5(-1.6 to -1.3) |
| Inflations | 3,521 | 3,703 |  | 3,291 | 4,065 |  |

\* $p<0.001$

**Table 3:** Anova Repeated Measures Adjusted Means Leak and tve 95%CI.

|  | TPR |  | SIB |  |
| --- | --- | --- | --- | --- |
|  | Blinded | Unblinded | Blinded | Unblinded |
| **Term** |  |  |  |  |
| tve | 20.3mL (20.2 - 20.5) | 23.2mL (23.0 - 23.4) | 29.4mL (29.2 – 29.7) | 28.3mL (28.1 to 28.6) |
| leak | 29.9% (29.3 to 30.6) | 16.4% (15.7 - 17.1) | 15.8% (15.3 – 16.3) | 10.5% (10.0 to 11.0) |
| **Preterm** |  |  |  |  |
| tve | 4.6mL (4.6-4.7) | 5.0mL (4.9 -5.1) | 3.7mL (3.7 - 3.8) | 4.6mL (4.5 - 4.6) |
| leak | 33.1% (32.4 - 33.7) | 18.4% (17.7 - 19.0) | 38.0% (37.4 - 38.7) | 22.6% (22.1 - 23.2) |

## DISCUSSION

The results from this study in a group of experienced NICU clinicians showed that using the Juno monitor significantly improved face mask leak and the targeted ventilation delivery in each manikin model using both resuscitation device types. Both resuscitation devices provide manual PPV lung inflation by notably different methods. The SIB provides inflation by inflation flow and pressure delivery that is dynamic, dependent on the user’s hand squeeze distance of the bag and has been shown to deliver more variable respiratory pressures and volume than TPR. ^15 16^ The TPR resuscitator is flow dependent, providing a preset PEEP and PIP more consistently^15 17^ but pressure alterations are less easily achieved during PPV. ^18^ The Neopuff manufacturer only recommends presetting of delivery pressures before use on a patient^19^, while NRP guidelines advise adjusting TPR PIP during PPV (MR SOPA) with the assistance of a second person^12^. The ability to dynamically adjust the PIP during resuscitation as a proxy measure for delivered volume is limited when using TPR compared to SIB (manometer fitted). Less experienced first responders using TPR systems may not factor in the need to increase PIP settings given a slow response of the patient when awaiting more experienced resuscitators to arrive and assist.

The limitations of this study are: 1. no subjects were in the “first responder” experience level, we speculate a greater benefit with first responder use; 2. no user adjustments of PIP when using TPR to increase tidal volumes (Figure 2) whereas with SIB, the volume was adjusted dynamically by squeeze distance and 3. it was not possible to tell if subjects used the tidal volume icon range or displayed volume as we did not use video recordings. The apparent ambiguity in instructions occurred as the study was designed to enroll an equal number of midwives as first responders, however, this was not possible during the Covid pandemic. As with most manikin studies the duration of PPV measured was brief. Sicker newborns often require a more prolonged duration of PPV. We speculate that fatigue and distraction may worsen mask leak and performance in this situation highlighting the need for dynamic monitoring. There were statistically significant but small differences in predicted mean tidal volumes within the accuracy tolerance of RFM’s pneumotach. This is explained by our overall low leak values in our experienced subjects and that the impact on reduced tve’s occurred mostly with leak values >60% (Figure 2).

Current ILCOR consensus on the use of RFM is based on devices not commercially available, ^4 8 20^ and are complex systems with GUI’s more suited to ICU and anesthetic domains than birthing environments where unexpected resuscitation is common. First responders have less experience than neonatal staff who may take valuable minutes to arrive on the scene. Even brief periods of over-ventilation can injure the preterm fetal lung and brain. ^21-23^ Relying on subjective assessment of chest wall movement may result in considerable variation in tidal volumes with under and over-ventilation.

The latest ILCOR statement on RFMs does not recommend their routine use during resuscitation^6^. The consensus of science knowledge gaps on the key research questions of the role of RFM use during newborn resuscitation include assessments of improving targeted ventilation and defining problematic average mask leak in general resuscitation delivery which our study provides insight.

This study shows experienced resuscitators with prior extensive training and competency in using the Juno monitor significantly improved mask seal and targeted ventilation in both extreme preterm and term manikin models. A study of 50 first responders is currently underway. Further research examines in detail clinicians’ experiences of the use of this monitor and, the effect on the duration of skills improvements are in planning. The first-in-human safety study of the clinical monitor version (Nemo) has recently been completed^24^.

## CONCLUSION

A new and novel in-line RFM monitor (Juno) displaying mask leak and deflation volume significantly reduced mask leak in a group of experienced resuscitators and reduced the number of high leak inflations in both term and preterm manikin models when used with SIB and TPR resuscitator systems.

## Data Availability

All data produced in the present study are available upon reasonable request to the authors

## Author Contribution statement

MT study design, data collection, data and statistical analysis, and writing of manuscript.

MH data collection, data and statistical analysis, and writing of manuscript.

MB and MC prototype ‘Juno’ monitor development and construction, data collection, and review of manuscript.

SM, KL, AP and MC data collection, manuscript writing, construction, and review.

