## Supplementary Figures for "Randomised study of a new inline respiratory function monitor (Juno) to improve mask seal and delivered ventilation with neonatal manikins"

Figure 3S: Current commercial Juno monitor display.

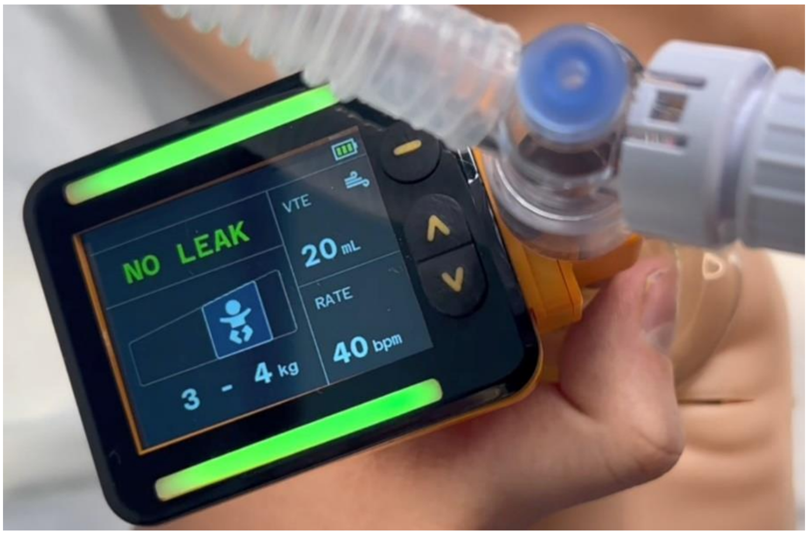

Figure 4S: icon graphic display of features pre-commercial prototype.

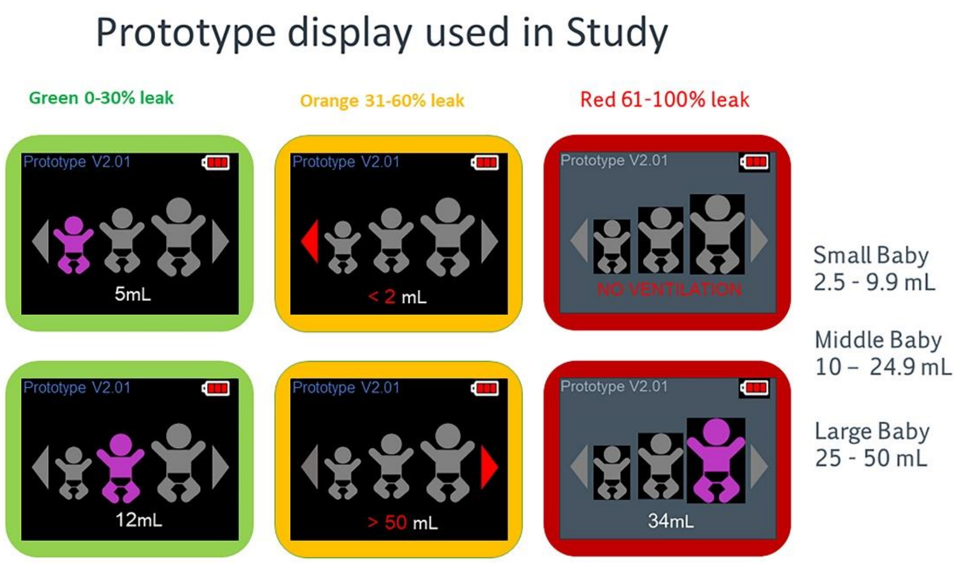
